# Diagnostic accuracy of a point-of-care urine tenofovir assay, and associations with HIV viraemia and drug resistance among people receiving dolutegravir and efavirenz-based antiretroviral therapy

**DOI:** 10.1101/2023.04.12.23288474

**Authors:** Jienchi Dorward, Richard Lessells, Katya Govender, Pravikrishnen Moodley, Natasha Samsunder, Yukteshwar Sookrajh, Phil Turner, Christopher C Butler, Gail Hayward, Monica Gandhi, Paul K. Drain, Nigel Garrett

**Author notes:** **Corresponding Author:** Jienchi Dorward, Nuffield Department of Primary Care Health Sciences, University of Oxford, Radcliffe Primary Care Building, Radcliffe Observatory Quarter, Woodstock Road, Oxford, OX2, 6GG.

## Abstract

**Introduction:** Novel point-of-care assays which measure urine tenofovir (TFV) concentrations may have a role in improving adherence monitoring for people living with HIV (PLHIV) receiving antiretroviral therapy (ART). However, further studies of their diagnostic accuracy, and whether results are associated with viraemia and drug resistance, are needed to guide their use, particularly in the context of the global dolutegravir rollout.

**Methods:** We conducted a cross-sectional evaluation among PLHIV receiving first-line ART containing tenofovir disoproxil fumarate (TDF). We calculated the diagnostic accuracy of the Abbott point-of-care assay to detect urine TFV measured by liquid chromatography and mass spectrometry. We evaluated the association between point-of-care urine TFV results and self-reported adherence, viraemia ≥1000 copies/mL, and HIV drug resistance, among people receiving either efavirenz or dolutegravir-based ART.

**Results:** Among 124 participants, 55% were women, median age was 39 (IQR 34-45) years. 74 (59.7%) were receiving efavirenz, and 50 (40.3%) dolutegravir. Sensitivity and specificity to detect urine TFV ≥1500ng/mL were 96.1% (95%CI 90.0-98.8) and 95.2% (75.3-100.0) respectively. Urine TFV results were associated with short (p<0.001) and medium term (p=0.036) self-reported adherence. Overall, 44/124 (35.5%) had viraemia, which was associated with undetectable TFV in those receiving efavirenz (OR 6.01, 1.27-39.0, p=0.014) and dolutegravir (OR 25.7, 4.20-294.8, p<0.001). However, in those with viraemia while receiving efavirenz, 8/27 (29.6%) had undetectable urine TFV, compared to 11/17 (64.7%) of those receiving dolutegravir. Drug resistance was detected in 23/27 (85.2%) of those receiving efavirenz and only 1/16 (6.3%) of those receiving dolutegravir. There was no association between urine TFV results and drug resistance.

**Conclusions:** Among PLHIV receiving ART, a rapid urine TFV assay can be used to accurately monitor urine TFV levels. Undetectable point-of-care urine TFV results were associated with viraemia, particularly among people receiving dolutegravir.

**Trial registration:** Pan-African Clinical Trials Registry: PACTR202001785886049.

## INTRODUCTION

Early identification and management of HIV viraemia among people living with HIV (PLHIV) receiving antiretroviral treatment (ART) is important to ensure rapid viral re-suppression, which in turn prevents HIV transmission, the development of HIV drug resistance (HIVDR), and morbidity and mortality [1]. Viraemia may be caused by inconsistent adherence to effective ART and/or HIVDR, but in low- and middle-income countries (LMICs), resistance and drug level testing are not widely available [2], making management of viraemia more difficult for clinicians.

Tenofovir disoproxil fumarate (TDF) is a key component of World Health Organization recommended ART and is used by over 95% of people receiving ART in LMICs [3, 4], thereby serving as a potential target for objective adherence monitoring. TDF is metabolised to tenofovir (TFV), which is converted intracellularly to TFV-diphosphate (TFV-DP). TFV has a short plasma half-life [5] and is excreted renally, meaning it is readily measured in urine. TFV-DP accumulates in red blood cells (RBCs) and peripheral blood mononuclear cells and has a longer half-life[6] than plasma TFV. Urine TFV concentrations have mainly been evaluated among people without HIV, who are using TDF as part of pre-exposure prophylaxis [7], showing an association with HIV seroconversion [8, 9]. However, measurement can require liquid chromatography tandem mass spectrometry (LC-MS/MS), which is not feasible in many settings. More recently, point-of-care urine TFV assays using antibody-based technology have been developed which can be readily used in clinical settings to accurately detect TDF adherence within the past 2-5 days [10].

While several clinical trials are planned or underway to evaluate the clinical impact of point-of-care urine TFV assays on HIV treatment outcomes[11, 12, 13], more observational data is needed to further evaluate diagnostic accuracy, and associations between point-of-care urine TFV results with HIV viraemia and drug resistance. In the context of the global rollout of the fixed-dose combination of tenofovir-lamivudine-dolutegravir (TLD), objective measures of adherence may be even more useful. Dolutegravir is an integrase inhibitor (INSTI) with a high genetic barrier to resistance and emergent drug resistance has so far been rare [14], meaning viraemia is most likely secondary to non-adherence. In contrast, previously recommended non-nucleoside reverse transcriptase inhibitors (such as efavirenz) are more susceptible to the development of drug resistance, meaning viraemia may be caused by either poor adherence or drug resistance. Thereby, demonstrating good adherence in the presence of viraemia on efavirenz based ART may help identify people with drug resistance.

In this study, we aimed to evaluate the diagnostic accuracy of a point-of-care test to detect urine TFV, compared to the reference standard LC-MS/MS. We also aimed to describe the association between point-of-care urine TFV results, and HIV viraemia, drug resistance, and self-reported adherence, among people receiving dolutegravir versus efavirenz. We hypothesized that a detectable urine TFV result on the point-of-care test would be more strongly associated with viral suppression among people receiving dolutegravir compared to efavirenz, because of the lower likelihood of drug resistance with dolutegravir. We also hypothesised that in people with confirmed viraemia, a detectable urine TFV result would be associated with HIV drug resistance among people receiving efavirenz.

## METHODS

### Study design

We conducted a prospective diagnostic accuracy sub-study within the POwER study. POwER is an open-label, individually randomised, feasibility study of point-of-care HIV viral load (VL) testing to enhance re-suppression among people with HIV viraemia while receiving first-line ART. The POwER protocol has been previously published[15].

### Participants

We included all consecutive participants enrolled into POwER who were taking tenofovir disoproxil fumarate. PLHIV were eligible for enrolment into POwER if they were receiving first-line dolutegravir or efavirenz-based ART and with recent viraemia >1000 copies/mL in the past 6 weeks, for which they had not yet received enhanced adherence counselling. At enrolment, participants provided sociodemographic and clinical details, including self-reported adherence, and had urine, dried blood spot and plasma samples taken and stored at -80°C, for retrospective testing.

### Test Methods

#### Point-of-care urine TFV testing

We thawed the frozen urine samples and tested them according to manufacturer’s instructions using the Abbott (Abbott, Chicago, USA) lateral flow point-of-care urine TFV assay. Specifically, 3-4 drops of urine were added to the test well, and the result was read by two independent laboratory technicians after 3-5 minutes. Photos of discrepant results were adjudicated by a third investigator.

#### Reference standard urine TFV, and TFV-DP concentrations

We used LC-MS/MS at the Africa Health Research Institute in Durban to quantitate TFV levels in thawed urine samples, and measure TFV-DP concentrations in dried blood spots (DBS) (see supplementary material for details).

#### Viral load and HIVDR

We tested VL with the cobas^®^ HIV-1 assay using the cobas 6800 platform (Roche, Basel, Switzerland) in the National Health Laboratory Service at the Inkosi Albert Luthuli Hospital in Durban. We attempted sequencing of HIV-1 *pol* (protease [PR], reverse transcriptase [RT] and integrase [IN]) for all samples with VL ≥1000 copies/mL. Following RNA extraction, we amplified PR, RT and IN genes using the amplification module of the Applied Biosystems HIV-1 Genotyping Kit with Integrase (Thermo Fisher Scientific, Waltham, MA); and sequenced on the Illumina MiSeq platform (Illumina, San Diego, CA). We identified drug resistance mutations (DRMs) at >20% frequency using Stanford HIVdb (version 9.1).

For all the above testing methods, the person conducting the test was blinded to the results from other methods.

### Statistical analysis

We assessed the diagnostic accuracy (sensitivity, specificity and positive and negative predictive values), of the point-of-care TFV test to detect urine TFV at the manufacturer stated threshold of 1500ng/mL. We compared self-reported short-term and longer-term adherence with point-of-care urine TFV results using logistic regression models. We then described the proportions of PLHIV with and without viraemia ≥1000 copies/mL who had detectable and undetectable point-of-care urine TFV tests. To determine the usefulness of the urine TFV test to predict viraemia and suppression in this study population, we calculated the pre-test probability (prevalence of viraemia and suppression before testing), and the post-test probability (prevalence after stratification by urine TFV test result). Among participants with a VL ≥1000 copies/mL, we also assessed proportions with and without HIV drug resistance who had detectable and undetectable point-of-care urine TFV results, and pre- and post-test probabilities for drug resistance. We conducted the above analyses separately among participants receiving efavirenz or dolutegravir. We also conducted sensitivity analyses using a viraemic threshold of ≥50 copies/mL. Lastly, among people with unexpected urine TFV results based on viraemia and HIV drug resistance results (e.g. viraemia, no HIV drug resistance, but *detectable* point-of-care urine TFV), we assessed longer term adherence by describing TFV-DP levels in DBS[6, 16]. Sample size was determined by the number of participants enrolled into POwER and receiving TDF. We analysed data using R 4.2.0 (R Foundation for Statistical Computing, Vienna, Austria).

### Ethical approvals

The eThekwini Municipality Health Unit Research Committee, the KwaZulu-Natal Provincial Health Research Ethics Committee (KZ_202002_005), the University of KwaZulu-Natal Biomedical Research Ethics Committee (BREC 00000836/2019) and the University of Oxford Tropical Research Ethics Committee (OxTREC 66-19) approved the study. POwER is registered on the Pan African Clinical Trials Registry (PACTR202001785886049) and the South African Clinical Trials Registry (DOH-27-072020-6890).

## RESULTS

### Study population

Between August 20^th^, 2020 and March 25^th^, 2022, we enrolled 125 participants into POwER (Figure 1), of whom 124 were receiving TDF and were included in this analysis. The median age was 39 years (interquartile range [IQR] 34-45) and 68 (54.8%) were women (Table 1). A total of 74 (59.7%) were receiving efavirenz for a median of 4.2 years (2.1 to 6.0), and 50 (40.3%) were receiving dolutegravir, for a median of 0.6 years (0.5-1.0). Median time since the pre-enrolment viraemic VL was 15 days (13-21). In December 2020, we were informed that 45 participants had their pre-enrolment VLs measured on a defective VL analyser which had been overestimating some VL results. Therefore, the viraemic sample used to determine eligibility may have been falsely high. After discussion with the ethics committees, we continued to include these participants in POwER and in this sub-study in order to have a wide range of VL results. At enrolment, 57/124 (46.0%) were virally suppressed <50 copies/mL, 23/124 (18.5%) had VL 50-999 copies/mL, and 44/124 participants (35.5%) had viraemia ≥1000 copies/mL. Among the 43 with successful HIVDR testing 24/43 (55.8%) had mutations conferring resistance to their current regimen. Among those receiving efavirenz, 23/27 (85.2%) had resistance to their current regimen, versus 1/16 (6.3%) of those receiving dolutegravir (one person with M184V mutation alone). Overall, 23.4% self-reported missing a dose in the past 4 days, and 62.9% reported last missing a dose over four weeks ago (**Error! Reference source not found**.).

**Figure 1:**
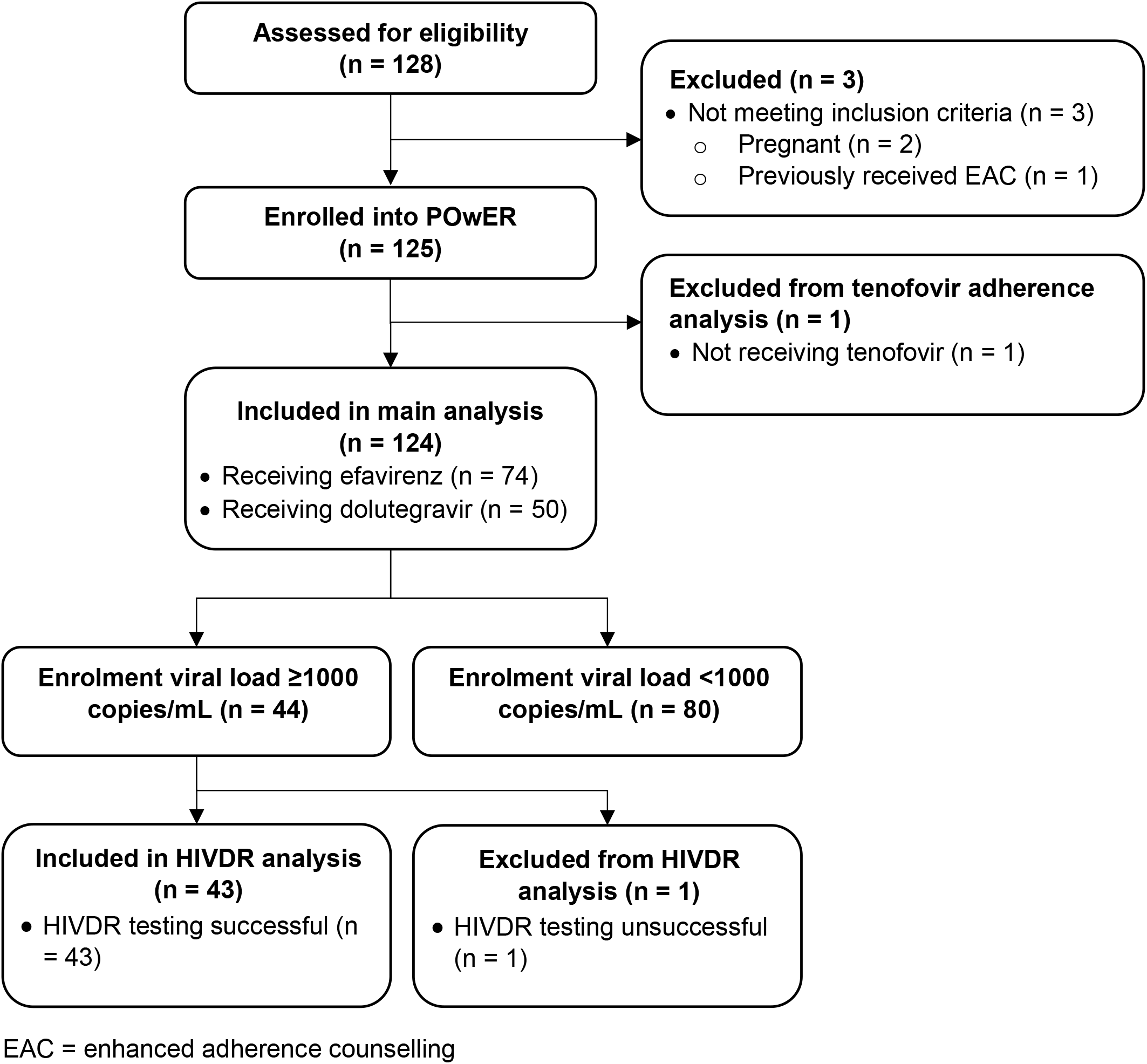
Flow diagram of POwER study participants.

**TABLE 1:** BASELINE DEMOGRAPHICS OF STUDY POPULATION, N = 124.

| Variable | Levels | Total |
| --- | --- | --- |
| Age, years | Median (IQR) | 39.0 (34.0 to 45.0) |
| Gender | Female | 68 (54.8) |
|  | Male | 56 (45.2) |
| Ethnicity | Black African | 121 (97.6) |
|  | Coloured | 1 (0.8) |
|  | Other | 2 (1.6) |
| Time since ART initiation, years | Median (IQR) | 4.1 (1.5 to 6.1) |
| Current ART regimen | TDF / FTC / EFV | 74 (59.7) |
|  | TDF / 3TC / DTG | 50 (40.3) |
| Time on current regimen, years | Median (IQR) | 1.6 (0.7 to 5.0) |
| ART side effects | No | 120 (96.8) |
|  | Yes | 4 (3.2) |
| Enrolment CD4 count category, cells/μL | <200 | 21 (16.9) |
|  | 200-349 | 25 (20.2) |
|  | 350-499 | 29 (23.4) |
| | ≥500 | 49 (39.5) |
| Last time participant missed a dose of ART | <2 weeks | 30 (24.2) |
|  | 2-4 weeks | 16 (12.9) |
|  | 1-3 months | 16 (12.9) |
|  | >3 months | 8 (6.5) |
|  | Never | 54 (43.5) |
| Number of ART doses missed in past 4 days | 0 | 95 (76.6) |
|  | 1 | 14 (11.3) |
|  | 2 | 9 (7.3) |
|  | 3 | 2 (1.6) |
|  | 4 | 4 (3.2) |
| POC urine tenofovir test | Not Present | 24 (19.4) |
|  | Present | 100 (80.6) |
| Urine tenofovir concentration, ng/mL | Median (IQR) | 20000.0 (7280.0 to 33625.0) |
| Enrolment viral load, copies/mL | <50 | 57 (46.0) |
|  | 50-999 | 23 (18.5) |
| | ≥1000 | 44 (35.5) |
| Any HIV drug resistance against current regimen? | No | 19 (15.3) |
|  | Yes | 24 (19.4) |
|  | Unsuccessful | 1 (0.8) |
|  | Viral load <1000 copies/mL | 80 (64.5) |

### Point-of-care urine TFV test

#### Diagnostic accuracy

At enrolment, 100 (80.6%) had urine TFV detected with the point-of-care test. Median LCMS-MS urine TFV concentration was 20000 ng/mL (7280-33625). Compared to quantitative urine TFV concentrations, the point-of-care TFV test was accurate at detecting TFV at the manufacturer stated threshold of 1500 ng/mL, with a sensitivity of 96.1% (95% confidence interval [CI] 90.0-98.8), and specificity of 95.2% (75.3-100.0, Table 2). All five discrepant results occurred in samples with LCMS-MS TFV between 500 and 3000ng/mL (Table S1).

**TABLE 2:** ANALYTIC PERFORMANCE OF THE POINT-OF-CARE TENOFOVIR TEST TO DETECT URINE TENOFOVIR AT THE MANUFACTURER THRESHOLD OF 1500NG/ML.

|  |  | LCSM urine TFV (ng/mL) |  |  |
| --- | --- | --- | --- | --- |
|  |  | <1500 | ≥1500 | Total |
| **POC TFV** | TFV not detected | 20 | 4 | 24 |
|  | TFV detected | 1 | 99 | 100 |
|  | Total | 21 | 103 | 124 |
| Sensitivity |  | 96.1% (90.0-98.8), p<0.001 |  |  |
| Specificity |  | 95.2% (75.3-100.0), p<0.001 |  |  |
| PPV |  | 99.0% (93.9-100.0), p<0.001 |  |  |
| NPV |  | 83.3% (63.4-93.8), p=0.002 |  |  |

#### Self-reported adherence

Among all participants, self-reported missed doses in the past four days and more recently self-reported missed doses were both associated with undetectable point-of-care urine TFV (Table S2). However, 12/95 (12.6%) people who reported missing no doses in the past four days had undetectable urine tenofovir. Two of these were false negatives, with LC-MS/MS concentrations >1500 ng/mL, and so were incorrectly classified as ‘non-adherent’ by the point-of-care test.

#### Association with viraemia

Undetectable TFV on point-of-care urine testing was associated with viraemia ≥1000 copies/mL in people receiving efavirenz (odds ratio [OR] 6.01, 1.27-39.0, p=0.014) and dolutegravir (OR 25.7, 4.20-294.8, p<0.001). Among those with viraemia ≥1000 copies/mL, 29.6% (13.8-50.2) of those receiving EFV had undetectable TFV, compared to 64.7% (41.1-82.7) of those receiving DTG (Fisher’s exact test for difference p=0.031, Table 3). Among those with viral suppression <1000 copies/mL, 93.6% (82.0-98.4) of those receiving EFV and 93.9% (79.2-99.2) of those receiving DTG had a detectable urine TFV.

**TABLE 3:**
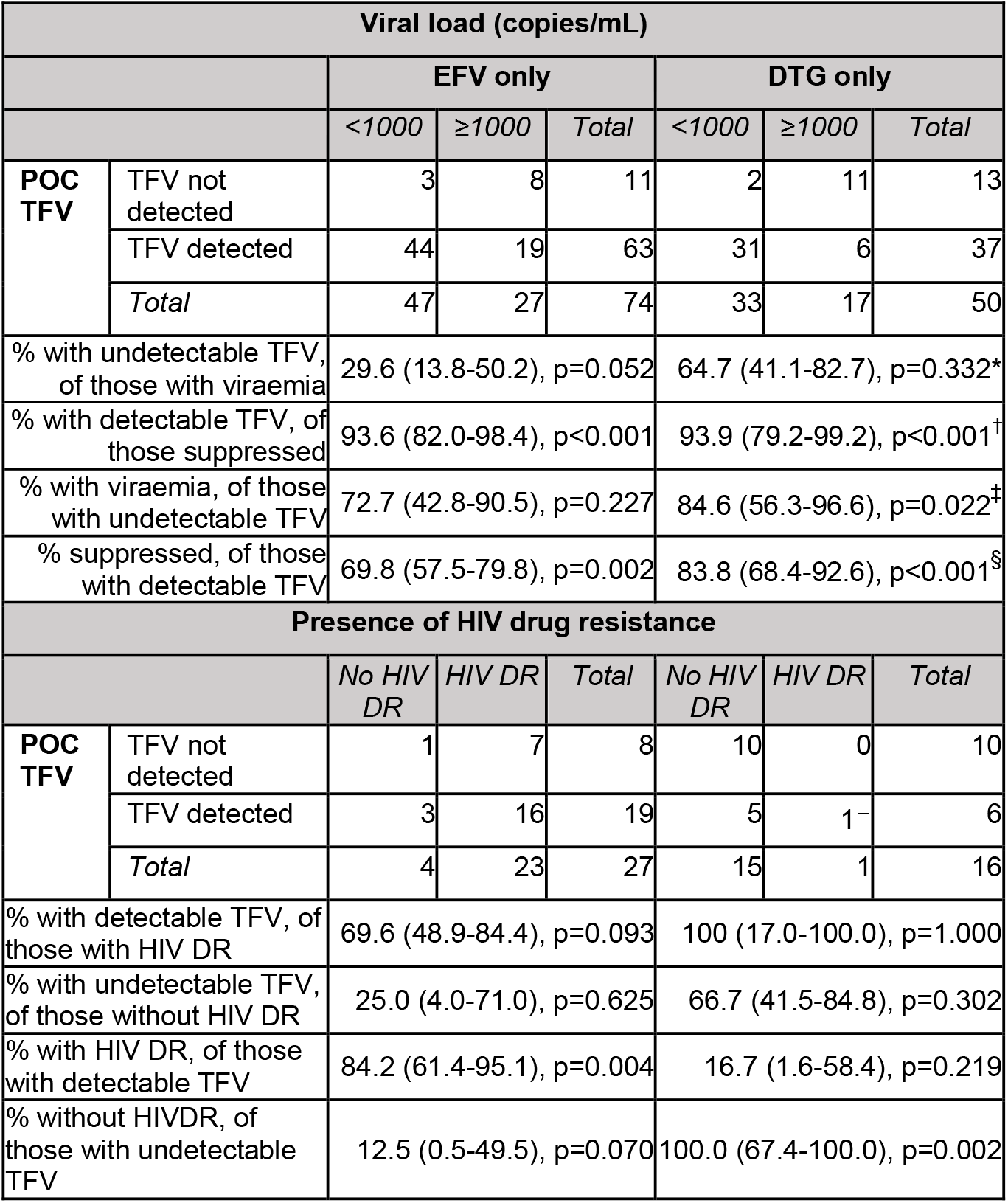
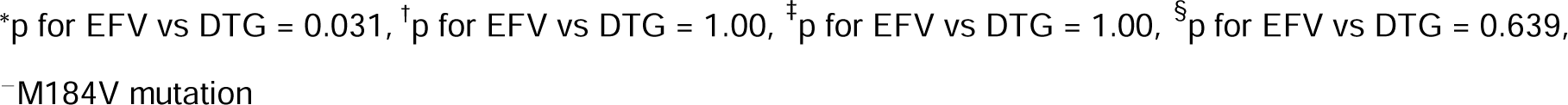
ASSOCIATION OF POINT-OF-CARE URINE TFV RESULTS WITH VIRAEMIA, AND HIV DRUG RESISTANCE.

| Viral load (copies/mL) |  |  |  |  |  |  |  |
| --- | --- | --- | --- | --- | --- | --- | --- |
|  |  | EFV only |  |  | DTG only |  |  |
|  |  | <1000 | ≥1000 | Total | <1000 | ≥1000 | Total |
| POC TFV | TFV not detected | 3 | 8 | 11 | 2 | 11 | 13 |
|  | TFV detected | 44 | 19 | 63 | 31 | 6 | 37 |
|  | Total | 47 | 27 | 74 | 33 | 17 | 50 |
| % with undetectable TFV, of those with viraemia |  | 29.6 (13.8-50.2), p=0.052 |  |  | 64.7 (41.1-82.7), p=0.332* |  |  |
| % with detectable TFV, of those suppressed |  | 93.6 (82.0-98.4), p<0.001 |  |  | 93.9 (79.2-99.2), p<0.001† |  |  |
| % with viraemia, of those with undetectable TFV |  | 72.7 (42.8-90.5), p=0.227 |  |  | 84.6 (56.3-96.6), p=0.022‡ |  |  |
| % suppressed, of those with detectable TFV |  | 69.8 (57.5-79.8), p=0.002 |  |  | 83.8 (68.4-92.6), p<0.001§ |  |  |
| Presence of HIV drug resistance |  |  |  |  |  |  |  |
|  |  | No HIV DR | HIV DR | Total | No HIV DR | HIV DR | Total |
| POC TFV | TFV not detected | 1 | 7 | 8 | 10 | 0 | 10 |
|  | TFV detected | 3 | 16 | 19 | 5 | 1 | 6 |
|  | Total | 4 | 23 | 27 | 15 | 1 | 16 |
| % with detectable TFV, of those with HIV DR |  | 69.6 (48.9-84.4), p=0.093 |  |  | 100 (17.0-100.0), p=1.000 |  |  |
| % with undetectable TFV, of those without HIV DR |  | 25.0 (4.0-71.0), p=0.625 |  |  | 66.7 (41.5-84.8), p=0.302 |  |  |
| % with HIV DR, of those with detectable TFV |  | 84.2 (61.4-95.1), p=0.004 |  |  | 16.7 (1.6-58.4), p=0.219 |  |  |
| % without HIVDR, of those with undetectable TFV |  | 12.5 (0.5-49.5), p=0.070 |  |  | 100.0 (67.4-100.0), p=0.002 |  |  |

\*p for EFV vs DTG = 0.031, <sup>†</sup>p for EFV vs DTG = 1.00, <sup>‡</sup>p for EFV vs DTG = 1.00, <sup>§</sup>p for EFV vs DTG = 0.639, <sup>–</sup>M184V mutation

Among all people receiving efavirenz, the pre-test probability was 36.5% (26.5-47.9) for viraemia and 63.5% (52.1-73.5) for viral suppression. After urine TFV testing, of those with undetectable urine TFV, the post-test probability was 72.7% (42.8-90.5) for viraemia, and of those with detectable urine TFV, 69.8% (57.5-79.8) for viral suppression (Table 3). Among those receiving dolutegravir, the pre-test probability was 34.0% (22.4-47.9) for viraemia and 66.0% (52.1-77.6) for viral suppression. Of those with undetectable urine TFV, the post-test probability was 84.6% (56.3-96.6) for viraemia, and of those with detectable urine TFV, 83.8% (68.4-92.6) for viral suppression.

At a threshold of ≥50 copies/mL, undetectable point-of-care urine TFV results remained associated with viraemia among those receiving efavirenz (p = 0.018) and those receiving dolutegravir (p = 0.002, Table S3), with 100% of those with undetectable TFV having viraemia.

#### Association with HIV drug resistance

Among 43 people with viraemia and successful HIV drug resistance testing, detectable point-of-care urine TFV results were not associated with HIV drug resistance against the current regimen in those receiving efavirenz (p = 1.000) or dolutegravir (p = 0.375). Of those receiving efavirenz, and with viraemia ≥1000 copies/ml, 23/27 (85.2%, 66.7-94.6) had drug resistance to their current regimen, and of these 23, 16 had a detectable urine TFV (69.6%, 48.9-84.4, Table 2). Of those without HIVDR, 1/4 (25.0%, 4.0-71.0) had undetectable urine TFV. Among those receiving dolutegravir, only 1/16 (6.3%, 0.0-30.6) had drug resistance to their current regimen (M184V mutation). This one participant had a detectable urine TFV test, while 10/15 without HIVDR (66.7%, 41.5-84.8) had undetectable urine TFV. When looking at only drug resistance against nucleoside reverse transcriptase inhibitors (NRTIs), there was again no association with urine TFV results among those receiving efavirenz (p=0.658) or dolutegravir (p=0.375).

Regarding the ability of the urine TFV test to predict HIVDR among people with viraemia, among people receiving efavirenz, the pre-test probability of HIVDR was 85.2% (66.7-94.6), and of not having HIVDR 14.8% (5.4-33.3). 19 people had detectable urine TFV, of whom 16 had HIVDR (post-test probability of HIVDR 84.2%, 61.4-95.1). Eight had undetectable urine TFV, and of these, only one did not have HIVDR (post-test probability of no HIVDR 12.5%, 0.5-49.5). Among people with viraemia and receiving dolutegravir, the pre-test probability of HIVDR was 6.3% (0.0-30.6), and of not having HIVDR, 93.7%. Six had detectable urine TFV, of which only one had HIVDR (post-test probability of HIVDR 16.7%, 1.6-58.4). Ten had undetectable TFV, and of these none had drug resistance (post-test probability of no HIVDR 100%, 67.4-100.0).

#### TFV-DP levels

Among the eight participants with viraemia, no HIVDR, and *detectable* urine TFV, one participant had LCMS urine TFV levels <1500ng/mL, suggesting a false positive, detectable point-of-care urine TFV test. Of the remaining seven, urine TFV levels were >1500ng/mL, suggesting recent TDF ingestion, but 6/7 had TFV-DP levels <700 fmol/punch, which corresponds to poor longer-term adherence with an estimated 0-3 tablets per week[6, 16] (Table S4A). Among the five participants with viral load <1000 copies/mL, but *undetectable* urine TFV, two had LMCS urine TFV levels >1500ng/mL, suggesting false negative, undetectable point-of-care urine TFV results. The remaining three participants had TFV-DP levels between 200 and 550 fmol/punch, again suggesting inconsistent longer-term adherence (Table S4B).

## DISCUSSION

### Summary

In this cross-sectional study in South Africa, we demonstrate that a novel point-of-care test is accurate at detecting urine TFV, and identifies additional people with sub-optimal adherence compared to self-reported adherence measures. Furthermore, undetectable point-of-care urine TFV results were associated with viraemia, in particular among people receiving dolutegravir. Among people with viraemia, point-of-care urine TFV results were not associated with HIV drug resistance.

### Interpretation

#### Self-reported adherence

We demonstrated an association between self-reported adherence and urine TFV levels, and found that around 10% of people who reported missing no doses in the past four days had undetectable urine tenofovir using the point-of-care test. Therefore, this test could help identify people with unreported sub-optimal adherence.

#### Viraemia

Undetectable urine TFV results were associated with viraemia, in particular among people receiving dolutegravir. Only 32% of those with viraemia while receiving efavirenz had an undetectable urine TFV; the majority had detectable urine TFV. However, this discrepancy can be explained by the high prevalence of HIV drug resistance among people receiving efavirenz, meaning that drug resistance, rather than current poor adherence, was driving viraemia.

Conversely, among people receiving dolutegravir, 63% of those with viraemia had undetectable urine TFV, suggesting that for these participants, poor adherence was the main cause of viraemia. This is supported by the low prevalence of HIV drug resistance in participants on dolutegravir. Furthermore, among those with viraemia and a detectable urine TFV, TFV-DP results suggested poor longer term adherence. In our study, the post-test probability of viraemia or suppression at a threshold of 1000 copies/mL was over 80% among those receiving dolutegravir, suggesting that this test could be used to triage people receiving dolutegravir into different clinical pathways. At a threshold of 50 copies/mL, an undetectable urine TFV very strongly predicted viraemia, but a detectable urine TFV performed less well at confirming viral suppression, likely because of the more sustained adherence required to suppress to <50 copies/mL.

#### HIV Drug Resistance

We did not find evidence of an association between urine TFV test results and HIV drug resistance among people with viraemia (≥1000 copies/mL) receiving efavirenz or dolutegravir, which may be partly explained by the small sample size, meaning that estimates were not precise. However, the very high prevalence of HIVDR among people receiving efavirenz (high pre-test probability of drug resistance), meant that the test did not ‘add value’, as the post-test probability of drug resistance remained similarly high. Likewise, the very low prevalence of resistance among people receiving dolutegravir (low pre-test probability), meant that the test was again not helpful, with post-test probabilities remaining similar.

### Comparison with other studies

There is one other published study assessing the analytic performance of the Abbot point-of-care tenofovir assay, conducted as part of its development and validation. In 300 randomly selected stored urine samples from the TARGET TDF dosing trial,[17] the point-of-care urine tenofovir assay had sensitivity of 97% (95-99) and specificity of 99% (94-100) at a cut-off of 1500ng/mL.[10] This cut-off was selected as it would correctly classify 98% of people who took a TDF dose 24 hours ago as adherent, and 86% of those who last took a dose 96 hours ago as non-adherent.[18] Of note, all the discrepant results in our analysis were close to the cut-off of 1500ng/mL.

Our findings are similar to those in recent studies which have assessed various point-of-care urine tenofovir assays, and their relationship with viraemia and/or HIVDR. A study from Lesotho among PLHIV receiving TDF-based ART (95% on dolutegravir) found that the UrSure point-of-care test (now known as SureQuick Rapid Tenofovir Adherence Test [OraSure Technologies Inc., USA]) did not detect urine TFV in 1/8 (12.5%) with viraemia ≥1000 copies/mL, and detected urine TFV in 395/398 (99%) of those with viral suppression [19]. Of the 8 with viraemia, 7 were receiving efavirenz-based ART, but HIVDR testing was not done.

A case control study within the ADVANCE clinical trial in South Africa matched 139 participants, recently initiated on ART and with rebound viraemia, with 53 non-viraemic controls [20]. 66% of participants with viraemia ≥200 copies/mL had an undetectable SureQuick Rapid Tenofovir Adherence Test, and 100% of those with viral suppression had a detectable urine TFV, which is similar to our findings. Among the 42 with successful drug resistance testing, drug resistance was detected in 16.7% of those receiving dolutegravir, and 61.1% of those receiving efavirenz. Overall, a detectable urine TFV result was associated with the presence of NRTI resistance alone, but not with the presence of NRTI/NNRTI resistance. This may reflect the persistence of NNRTI mutations in the absence of drug pressure, whereas NRTI mutations are more likely to be superceded by wild type virus if ART is not being taken.

Another South African study, among 113 people with previous viraemia or treatment interruptions, found that among people with viraemia ≥400 copies/mL, 64.7% of those receiving dolutegravir/boosted protease inhibitors had an undetectable Abbott point-of-care urine TFV result, compared to only 4.8% among those receiving efavirenz. Only one person receiving efavirenz had undetectable urine TFV [21]. Similar to our findings, among those receiving dolutegravir/boosted protease inhibitors, 85% of those with undetectable urine TFV had viraemia, and 85% of those with detectable urine TFV were suppressed. As in our study, the prevalence of HIVDR was high among people with efavirenz (18/20, 90%), and much lower among those receiving dolutegravir (5/16 = 31%), although associations between HIVDR and urine TFV results were not evaluated. Two other studies, focusing on people with viraemia while receiving efavirenz, found associations between positive point-of-care urine TFV results and HIVDR mutations, but similar to our findings, the prevalence of HIVDR mutations was high, and so the pre- and post-test probabilities of HIVDR did not change much after a positive urine TFV test[22, 23].

### Strengths and limitations

This is one of the first evaluations of the analytic performance of the Abbot point-of-care urine tenofovir assay, beyond the validation conducted as part of the assay’s development.[10] We were also able to use viral load and HIV drug resistance results to evaluate the clinical utility of the assay, and our inclusion of people receiving dolutegravir is important given the ongoing dolutegravir rollout. Limitations of our study include the use of frozen urine samples for retrospective point-of-care testing by laboratory staff, the short period which participants had been receiving dolutegravir, and the relatively small sample size, particularly of people with viraemia and drug resistance, which resulted in wide confidence intervals. Our study design meant that we enrolled people a median of 15 days after first viraemia, meaning that adherence patterns could have changed, particularly if people had collected ART at the visit when blood was taken for the first viraemic VL. This cross-sectional analysis also does not explore the relationship between current adherence measures and future outcomes.

### Implications for research and policy

Our findings provide further evidence to support the use of point-of-care urine TFV assays as objective indicators of adherence, and provide further insights into where they may be best employed in clinical practice. The moderate performance at predicting viraemia seen in our and other studies suggests that current point-of-care urine TFV assays cannot replace VL testing, but could be used in-between annual VL testing to monitor adherence, and to trigger VL testing if poor adherence is suspected. This may be particularly useful in people with viraemia receiving dolutegravir, or in differentiated ART delivery programmes in community settings. While our findings suggest limited utility in identifying resistance among people with viraemia while the prevalence of dolutegravir resistance is low, further work is needed to determine whether the point-of-care urine TFV test could be useful as a triage test to guide HIV drug resistance testing as the dolutegravir rollout matures.

## Conclusion

Point-of-care urine TFV may be used to monitor ART adherence and predict viraemia, particularly in people receiving dolutegravir.

## Data Availability

Bona fide researchers will be able to request access to anonymised trial data by contacting the corresponding author.

## LIST OF ABBREVIATIONS

ART: antiretroviral therapy
AUC: area under the curve
DBS: dried blood spot
HIVDR: HIV drug resistance
INSTI: integrase inhibitor
LMIC: low- and middle-income countries
NRTI: nucleotide reverse transcriptase inhibitor
NNRTI: non-nucleotide reverse transcriptase inhibitor
NPV: negative predictive value
PLHIV: people living with HIV
PPV: positive predictive value
ROC: receiver operating characteristic
TDF: tenofovir disoproxil fumarate
TFV: tenofovir
TFV-DP: tenofovir diphosphate
VL: viral load

## DECLARATIONS

### Author contributions

JD, PKD and NG conceived the study. RL, KG, PM and NS were responsible for laboratory testing. RL, YS, CB and GH contributed to study design. MG developed the urine TFV assay. JD analysed the data and wrote the first draft of the manuscript. All authors critically reviewed and edited the manuscript and consented to final publication.

## Acknowledgements

The authors would like to thank all participants in the study and acknowledge the work and support of staff at the Prince Cyril Zulu Clinic, Mafakathini Clinic, eThekwini Municipality, CAPRISA, AHRI Pharmacology Core, KRISP and the National Health Laboratory Services at Inkosi Albert Luthuli Hospital.

