## Supplement for "Diagnostic accuracy of a point-of-care urine tenofovir assay, and associations with HIV viraemia and drug resistance among people receiving dolutegravir and efavirenz-based antiretroviral therapy"

**SUPPLEMENTARY FILE**

### 1. LC-MS/MS sample analysis protocol

A quantitative LC-MS/MS method was developed for the determination of tenofovir (TFV) in urine samples and TFV and tenofovir-diphosphate (TFV-DP) concentrations in dry blood spot (DBS) samples. The LC-MS/MS method was accurate, robust and quantitative over the concentration ranges; 0.5 – 80 µg/mL for TFV in urine and 100 – 8000 pg/mL for TFV and TFV-DP in DBS samples.

The urine and DBS samples were processed using a protein precipitation method. A 70% methanol:water (v/v) solution, which contained the deuterated internal standards; d6-TFV and d5-TFV-DP was used for drug analyte extraction. The calibration standards and quality control solutions (containing TFV and TFV-DP) were prepared using the extraction solution.

The LC-MS/MS analysis was performed using an Agilent high pressure liquid chromatography (HPLC) system coupled to an AB Sciex 5500, triple quadrupole mass spectrometer equipped with an electrospray ionization (ESI) TurboIonSpray source. Analyst software, version 1.6.2 was used for data acquisition and quantitative data analysis.

Tenofovir and TFV-DP was quantitated using ion pair-hydrophilic interaction chromatography coupled to tandem mass spectrometry (IP–HILIC–MS/MS). The chromatographic separation was performed at a flow rate of 0.2 mL/min on a Luna Amino (NH2) column (Phenomenex, Torrance, CA) 100 mm × 2.0 mm, packed with 3.0 µm particles. Mobile phase A consisted of 100 mM hexafluoro-2-propanol (HFIP) and 0.5% diethylamine (DEA) (v/v) in water, and mobile phase B consisted of 0.1 M HFIP and 0.5% DEA (v/v) in acetonitrile. A sample volume of 5.0 µL was injected onto the HPLC column and the analytes were separated using a gradient elution. The autosampler syringe and the injection valve were washed with a water:acetonitrile (30:70, v/v) solution, post sample injection, to reduce carryover. The system was operated in negative-ion multiple reaction monitoring (MRM) mode set to detect precursor [M+H]^+^→ product ion transitions for TFV1 (*m/z* 285.8 → *m/z* 133.9), TFV2 (*m/z* 285.8→ *m/z* 151.0), TFV-DP1 (*m/z* 445.8 → *m/z* 158.9), TFV-DP2(*m/z* 445.8→ *m/z* 176.7) and the internal standard; d6-TFV (*m/z* 292.0→ *m/z* 133.8) and d5-TFV-DP (*m/z* 450.8→ *m/z* 158.9). The optimized ESI source dependent parameters were set as follows; ion spray voltage (ISV): 5500V, temperature (TEM): 350°C, gas 1 (N_2_) and gas 2 (N_2_): 40 psi.

### 2. Table S1: Point-of-care urine TFV results for samples measured between 500- 3000ng/mL with LCMS-MS

| **Participant** | **LCMS-MS Urine TFV concentration (ng/mL)** | **Point-of-care urine TFV result** | **Discrepant result?** |
| --- | --- | --- | --- |
| A | 613 | Present | Discrepant |
| B | 687 | Not Present | Not Discrepant |
| C | 697 | Not Present | Not Discrepant |
| D | 944 | Not Present | Not Discrepant |
| E | 1030 | Not Present | Not Discrepant |
| F | 1150 | Not Present | Not Discrepant |
| G | 1620 | Not Present | Discrepant |
| H | 2090 | Not Present | Discrepant |
| I | 2430 | Not Present | Discrepant |
| J | 2790 | Not Present | Discrepant |

LCMS-MS = Liquid chromatography tandem mass spectrometry

### 3. Table S2: Self-reported adherence compared to point-of-care urine TFV results

| **Adherence variable** | | **POC TFV assay** | | |
| --- | --- | --- | --- | --- |
|  |  | **Not Present** | **Present** | **Odds ratio* (95% CI), P** |
| Number of ART doses missed in past 4 days | 0 | 12 (12.6) | 84 (88.4) | 0.44 (0.27-0.67)  P = <0.001 |
|  | 1 | 3 (21.4) | 11 (78.6) |  |
|  | 2 | 5 (55.6) | 4 (44.4) |  |
|  | 3 | 1 (50.0) | 1 (50.0) |  |
|  | 4 | 3 (75.0) | 1 (25.0) |  |
| Last time participant missed a dose of ART (weeks) | Never | 5 (9.3) | 50 (92.6) | 0.66 (0.49-0.87)  P = 0.036 |
|  | >12 | 1 (12.5) | 7 (87.5) |  |
|  | 4-12 | 4 (25.0) | 12 (75.0) |  |
|  | 2-4 | 3 (18.8) | 13 (81.2) |  |
|  | <2 | 11 (36.7) | 19 (63.3) |  |

*Binomial logistic regression models.

POC = point-of-care, TFV = tenofovir

### 4. Table S3: Analytic performance of the point-of-care urine tenofovir test to detect viraemia ≥50 copies/mL

| **Viral load (copies/mL)** | | | | | | | | |
| --- | --- | --- | --- | --- | --- | --- | --- | --- |
|  | | | **EFV only** | | | **DTG only** | | |
|  | | *<50* | | *≥50* | *Total* | *<50* | *≥50* | *Total* |
| **POC TFV** | TFV not detected | 2 | | 9 | 11 | 0 | 13 | 13 |
|  | TFV detected | 38 | | 25 | 63 | 17 | 20 | 37 |
|  | *Total* | 40 | | 34 | 74 | 17 | 33 | 50 |
| % with undetectable TFV, of those with viraemia | | 26.5 (14.5-43.4), p=0.009 | | | | 39.4 (24.7-56.4), p=0.296* | | |
| % with detectable TFV, of those suppressed | | 95.0 (82.4-99.4), p<0.001 | | | | 100 (77.9-100), p<0.001^†^ | | |
| % with viraemia, of those with undetectable TFV | | 81.2 (51.0-95.7), p=0.065 | | | | 100 (72.9-100), p<0.001^‡^ | | |
| % suppressed, of those with detectable TFV | | 60.3 (48.0-71.4), p=0.130 | | | | 45.9 (31.1-61.6), p=0.743^§^ | | |

*p for EFV vs DTG = 0.469, ^†^p for EFV vs DTG = 1.00, ^‡^p for EFV vs DTG = 0.774, ^§^p for EFV vs DTG = 0.485

**Table S4: Viral load, HIV drug resistance, urine TFV and TFV-DP results for participants with unexpected point-of-care tenofovir results**

| **ID** | **ART regimen** | **Viral load (cps/mL)** | **Drug resistance against current ART?** | **POC TFV result** | **Quantitative urine TFV (ng/mL)** | **Quantitative DBS TFV-DP (fmol/punch)** | **Comment** |
| --- | --- | --- | --- | --- | --- | --- | --- |
| A) **Viraemia ≥1000 copies/mL** **with no drug resistance, but detectable POC urine TFV** | | | | | | | |
| 1 | TDF / FTC / EFV | 3550 | No | Detected | 26900 | 828 |  |
| 2 | TDF / FTC / EFV | 2570 | No | Detected | 62000 | 634 | Low TFV-DP |
| 3 | TDF / FTC / EFV | 1450 | No | Detected | 4080 | 219 | Low TFV-DP |
| 4 | TDF / 3TC / DTG | 4900 | No | Detected | 61200 | 90 | Low TFV-DP |
| 5 | TDF / 3TC / DTG | 2950 | No | Detected | 43100 | 477 | Low TFV-DP |
| 6 | TDF / 3TC / DTG | 11700 | No | Detected | 33800 | 573 | Low TFV-DP |
| 7 | TDF / 3TC / DTG | 14800 | No | Detected | 15000 | 0 | Low TFV-DP |
| 8 | TDF / 3TC / DTG | 3240 | No | Detected | 613 | 103 | False positive POC TFV |
| B) **Viral suppression <1000 copies/mL but undetectable POC urine TFV** | | | | | | | |
| 9 | TDF / FTC / EFV | <50 | NA | Not Detected | 0 | 201 | Low TFV-DP |
| 10 | TDF / FTC / EFV | <50 | NA | Not Detected | 1620 | 821 | False negative POC TFV |
| 11 | TDF / FTC / EFV | 340 | NA | Not Detected | 2090 | 483 | False negative POC TFV |
| 12 | TDF / 3TC / DTG | 90 | NA | Not Detected | 236 | 522 | Low TFV-DP |
| 13 | TDF / 3TC / DTG | 112 | NA | Not Detected | 252 | 354 | Low TFV-DP |

ART = antiretroviral therapy, POC = point-of-care, TFV = tenofovir, DBS = dried blood spot, TFV-DP = tenofovir diphosphate
